# SARS-CoV-2 antibody profile of naturally infected and vaccinated individuals detected using qualitative, semi-quantitative and multiplex immunoassays

**DOI:** 10.1101/2022.01.24.22269699

**Authors:** Jamie Meyers, Anne Windau, Christine Schmotzer, Elie Saade, Jaime Noguez, Lisa Stempak, Xiaochun Zhang

## Abstract

Antibody profiling of vaccinated versus naturally infected individuals is important in evaluating immunity and aiding diagnosis and booster strategies. This study measured antibodies against different antigen targets in healthcare workers (HCW) who have been fully vaccinated with mRNA vaccines, recovered from natural infection, or patients during active infection. All vaccinated individuals were positive for anti-RBD, anti-S1, and anti-S2 antibodies. The median index and interquartile range (IQR) by the Atellica IgG assay were 179 and 140-291 respectively. Among the different antigen targets within the Bioplex assay, the levels of anti-RBD were highest (median >3200 U/mL; IQR >3200 to >3200 U/mL), followed by anti-S1 (median 2132 U/mL; IQR 1649->3200 U/mL) and anti-S2 (median=42 U/mL; IQR=24-76 U/mL). The non-vaccinated recovered cohort showed 90% seropositivity (median index >10; IQR 3.6->10) by Atellica total antibody, 73% by Atellica IgG (median index 3.3; IQR 0.9-9.4), 84% by Bioplex anti-RBD (median 62; IQR 12-269 U/mL), 77% by Bioplex anti-S1 (median 45; IQR 10-83 U/mL), 37% by Bioplex anti-S2 (median 7; IQR 1-12 U/mL), and 79% by Bioplex anti-nucleocapsid (median 69; IQR 20-185U/mL) respectively. The active infection cohort exhibited a similar pattern as the recovered cohort. About 88% and 78% of the recovered and active infection cohort produced both anti-spike and anti-N antibodies with Anti-S1/anti-N ratios ranging from 0.07-16.26. In summary, fully vaccinated individuals demonstrated an average of 50-fold higher antibody levels than naturally infected unvaccinated individuals with immune reactivity strongly towards RBD/S1 and a weak response to S2. The results support vaccination regardless of previous COVID-infection status.

## Introduction

Immunity plays a critical role in preventing SARS-CoV-2 infection and protecting from severe disease. Vaccines have been proved to be powerful tools in fighting the pandemic (Fowlkes et al. 2021; Lopez Bernal et al. 2021; Sara Y Tartof 2021). Multiple large-scale studies with more than 10,000 participants also demonstrated that natural infection can induce an immune response that confers high levels of protection against symptomatic illness for at least 6 months (Hall et al. 2021; Sheehan et al. 2021; Vitale et al. 2021). Studies that compared protective immunity acquired from a previous infection versus vaccination showed controversial results. Both equal protection and greater protection of vaccines have been reported (Bozio et al. 2021; Shenai et al. 2021). Public health policies addressing whether or not to vaccinate people who have already been infected vary by country. For example, Israel allows people who have recovered from COVID-19 to get a vaccination “Green Pass” valid for up to six months, while the United States has a straightforward approach with only two categories, vaccinated or unvaccinated, regardless of a previous infection.

Circulating anti-SARS-CoV-2 antibody levels reflect the degree of the humoral immune response, an important part of adaptive immunity. Serologic tests measuring antibodies against various SARS-CoV-2 antigen targets are now widely available. Both qualitative and semi-quantitative antibody assays have been granted Emergency Use Authorization (EUA) by the FDA and utilized at clinical laboratories. So far, numerous publications reported antibody response due to vaccination, or after natural infection separately. However, direct comparisons have been scarce; due to the diverse antigen targets, different measurement units, and lack of traceability in calibration, comparison between vaccine-elicited and infection-induced antibody production across different studies/assays is not easy and can be misleading.

With more people vaccinated against SARS-CoV-2 and the increasing number of infections due to new variants, knowledge regarding the antibody profile of vaccinated versus naturally infected individuals is important in interpreting clinical lab antibody results and providing information for improving vaccine booster strategies. This study examined antibodies against spike protein (S) receptor-binding domain (RBD), S1, S2, and nucleocapsid (N) protein in healthcare workers (HCW) who have been fully vaccinated with mRNA vaccine, recovered from natural infection, or patients during active infection. It provides a direct comparison between vaccine- and infection-induced humoral immunity using two qualitative and two semi-quantitative immunoassays.

## Materials and Methods

### SUBJECTS AND SAMPLES

Residual blood samples from research studies and clinical testing at University Hospitals Cleveland Medical Center were used for the study. Three cohorts were included: fully vaccinated HCW, nonvaccinated HCW who recovered from natural infection, and patients with acute infection. The vaccination cohort was comprised of 33 healthcare worker volunteers who had received two doses of the Moderna mRNA vaccine. Samples were collected between 14 days to 1 month after the 2nd dose. The recoveredgroup included 61 healthcare workers who participated in a serosurveillance research study (IRB number: STUDY20200608) and were identified as past SARS-CoV-2 infection by either a positive result (n=52) of the IgG anti-N protein assay (Abbott, IL) or negative serology, but with a self-reported positive PCR result. The active infection group included consecutively collected symptomatic COVID patients diagnosed using PCR testing at University Hospitals (n=52 samples from 32 patients collected at various post symptom onset dates). All samples were utilized according to University Hospitals Institutional Review Board policies governing the use of residual material for assay validation.

### MEASURE SARS-COV-2 ANTIBODIES USING FOUR DIFFERENT ASSAYS

All assays were performed and interpreted per the manufacturer’s instructions. Bioplex 2200 semi-quantitative SARS-CoV-2 IgG multiplex panel (Bio-Rad, CA) detects and differentiates antibodies against the RBD, S1, S2, and nucleocapsid protein. The assay cutoff for positivity is 10 U/mL. Architect qualitative SARS-CoV-2 IgG assay detects Ig antibodies against nucleocapsid (Abbott, IL) with a cutoff of 1.4 index value. The Atellica SARS-CoV-2 semi-quantitative IgG assay and qualitative total antibody assay measure anti-RBD IgG and total antibodies, respectively (Siemens, NY) with a 1.0 index value as assay cutoff for positivity. Additional details regarding these assays were described in the supplemental Table S1.

### STATISTICAL METHODS

Statistical analyses were performed using SigmaPlot software. The difference between two groups was compared using the Student t test or Mann-Whitney Rank Sum Test depending on whether the Normality Test (Shapiro-Wilk) passed or failed. The difference among three or more groups was compared using the One-way ANOVA or Kruskal-Wallis One-way ANOVA on ranks depending on the result of the Normality Test. Spearman Rank Order Correlation was used to assess the correlation between variables such as seroprevalence and exposure to COVID-19. A p value <0.05 was regarded as statistically significant.

## Results

The basic characteristics of the specimens from the study cohorts are summarized in Table 1. The recovery cohort includes a wide spectrum of disease severity ranging from asymptomatic infection to hospitalization and ICU stay among healthcare workers before COVID-19 vaccines were available. The acute infection cohort represents an older and sicker population treated at our health system at the beginning of the pandemic.

**Table 1.** Demographic and clinical characteristics of study cohorts.

| **Table 1. Demographic and clinical characteristics of study cohorts** |  |
| --- | --- |
| **Fully vaccinated healthcare workers (HCW) (n=33)** |  |
| Vaccine | Moderna COVID-19 mRNA vaccine (2 shots, 28 days apart) |
| Age | Median: 42; Range: 23-63 years old |
| Gender | Female: 20; Male: 13 |
| Specimen collection time | Median: 16 days; Range: 14-30 days after 2nd dose |
| Previous COVID infection | 3 showed positive anti-N IgG confirmed by a second assay |
| **HCW recovered from natural COVID-19 infection** |  |
| (61 unvaccinated individuals infected between January and November 2020) |  |
| Age | Median: 41; Range: 23-66 years old |
| Gender | Female: 51; Male: 10 |
| Specimen collection time | Median: 134; Range: 17-249 days after symptom onset or positive PCR test |
| COVID PCR Test | 26 (42%) positive; 8 (13%) negative; 28 (45%) not tested |
| Symptom | 37 (62%) symptomatic; 23 (38%) asymptomatic |
| Hospitalization/ICU | 3 (5%) hospitalized and 1(2%) with ICU stay |
| **Acute active COVID-19 infection** |  |
| (32 symptomatic and PCR positive patients between March and May, 2020) |  |
| Age | Median: 60; Range: 24-83 years old |
| Gender | Female: 15; Male: 17 |
| Specimen collection time | multiple time points. Median: 8; Range: 3-25 days after symptom onset |
| Hospitalization/ICU | 24 (75%) hospitalized and 10 (31%) with ICU stay |

Antibody levels among individuals in the 2-dose vaccinated and non-vaccinated recovered cohort are shown in Tables 2 and 3, respectively. All the vaccinated individuals were positive by the S1 and RBD antibody assays with values exceeding the assay upper limit. For the semi-quantitative assays, all samples triggered auto-dilution and were further tested with appropriate dilutions per the assay manufacturer’s instructions. The median was approximately 261-,>320-, and 213fold of the cutoff levels of the Atellica IgG anti-RBD, Bioplex anti-RBD, and anti-S1 IgG assay, respectively. Even though 100% of the vaccinated individuals were positive for anti-S2 IgG, the S2 antibody levels were much lower than anti-S1 or anti-RBD, with the median being only about 4-fold of the cut-off value. Additionally, three individuals were also positive for anti-N IgG indicating previous natural infection. These three samples were the top 3 highest samples by the S1, RBD and S2 semi-quantitative assays. Anti-RBD values of these three samples were 3.5, 4.0, and 5.7 times the median level by the Siemens IgG assay. Interestingly, even though the response to S2 was generally much lower than S1, notable heterogeneity was observed. The three previously infected and vaccinated people had anti-S2 IgG levels ranging from 18 to >80 times the cutoff level, which was significantly higher than the naïve vaccinated people (p<0.05).

**Table 2.** Antibody levels of vaccinated individuals by multiple assays with different antigen targets.

|  | Total anti-RBD<br>(Siemens Atellica) | IgG anti-RBD<br>(Siemens Atellica) | IgG anti-RBD<br>(Biorad Bioplex) | IgG anti-S1<br>(Biorad Bioplex) | IgG anti-S2<br>(Biorad Bioplex) | IgG anti-N<br>(Biorad Bioplex) |
| --- | --- | --- | --- | --- | --- | --- |
| Positivity<br>(Positive/Total) % | (33/33)100% | (33/33)100% | (33/33)100% | (33/33)100% | (33/33)100% | (3/33)9% |
| Median index (or<br>unit) value | >10 | 261 | >3200 U | 2132 U | 42 U | <1U |
| Interquartile range<br>(IQR) | >10 ->10 | 140 - 291 | >3200 - >3200U | 1649 -> 3200U | 24 - 76U | <1 - <1U |

**Table 3.**
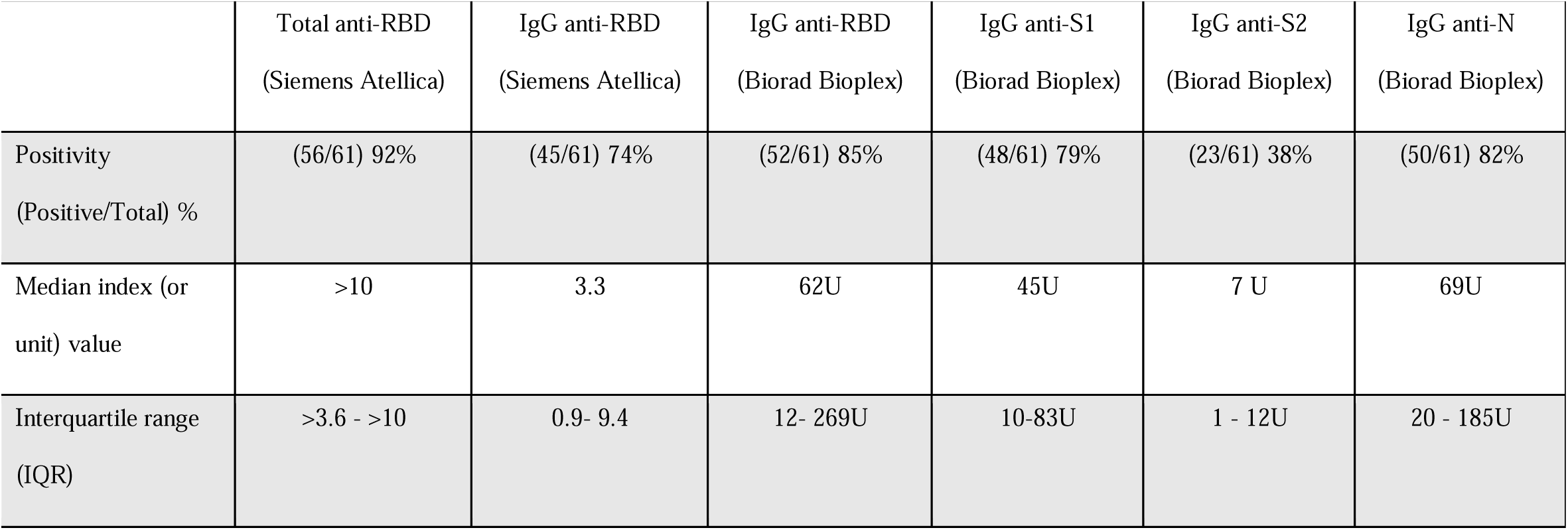
Antibody levels of unvaccinated individuals recovered from COVID-19 by multiple assays with different antigen targets.

Antibody results of unvaccinated individuals recovered from natural infection are summarized in Table 3. Seropositivity rates were different depending on antigen targets and assays. The total anti-RBD assay showed the highest positive rate of 90%, while the anti-S2 assay exhibited the lowest seropositivity at 37%. The median antibody levels of this cohort were 3, 6, and 4.5 times the assay cutoff by the Atellica anti-RBD, Bioplex anti-RBD, and anti-S1 IgG assay, respectively. About 2%, 40%, 22%, and 30% of samples exceeded the assay analytical range and needed further dilutions for analysis on the Atellica anti-RBD IgG, Bioplex anti-RBD, anti-S1, and anti-N IgG semi-quantitative assays, respectively.

As shown in Figure 1, anti-RBD, anti-S1, and anti-S2 antibody levels measured using the Bioplex assay were dramatically higher in the vaccinated individuals than the unvaccinated individuals recovered from natural infection. It was about 47- and 6-fold higher for S1 and S2, respectively. However, there’s no significant difference between the recovered and active infection cohorts. The results from the Atellica semi-quantitative anti-RBD IgG assay showed a similar pattern. The median in the vaccinated cohort was 54 times higher than the recovered cohort. Both the recovered and active infection cohorts presented a wide range of antibody levels. Two out of 32 patients (6%) with active infection had anti-RBD IgG greater than 179 (the median anti-RBD IgG index of the vaccinated cohort).

**Fig 1.**
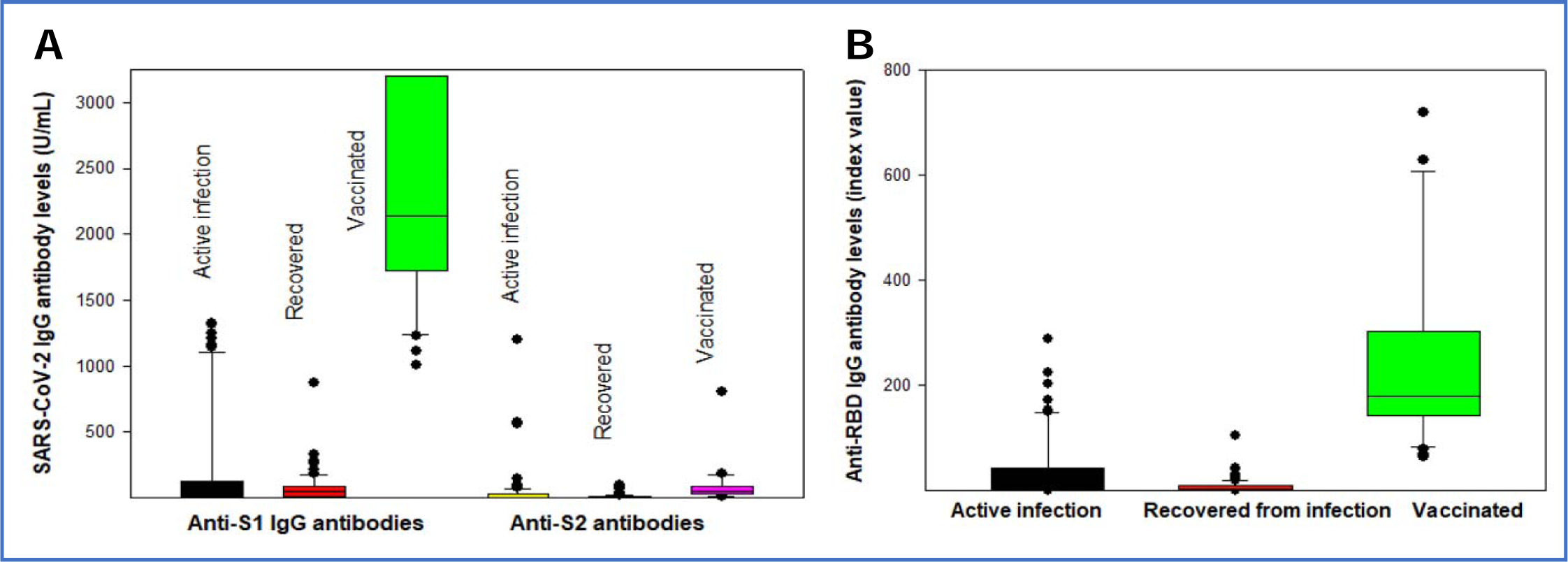
Comparison of SARS-CoV-2 antibody levels to different antigens between actively infected, recovered, and vaccinated cohorts. IgG antibody levels against S1 and S2 were significantly higher (about 47 and 6 folds, respectively) in vaccinated individuals than unvaccinated people who have recovered from natural SARS-CoV-2 infection (P<0.01). No significant difference between recovered and active infection in S1 or S2 antibodies.

We further analyzed the ratio of anti-S1 and anti-N IgG in each individual with active or past COVID infection using the multiplex assay data. As shown in Figure 2, most individuals (>78%) developed both anti-S1 and anti-N antibodies after natural infection. However, the anti-S1/anti-N ratio varied greatly between individuals. Additionally, a small but significant portion of the infected population produced only one type of antibody: either anti-N or anti-S. Further analysis of PCR Ct values and anti-S1/anti-N ratio in the active infection cohort did not show any correlation. In both the active and recovered infection cohorts, no significance of anti-S1/anti-N ratio was observed between outpatients, inpatients, and ICU patients (p>0.05).

**Fig 2.**
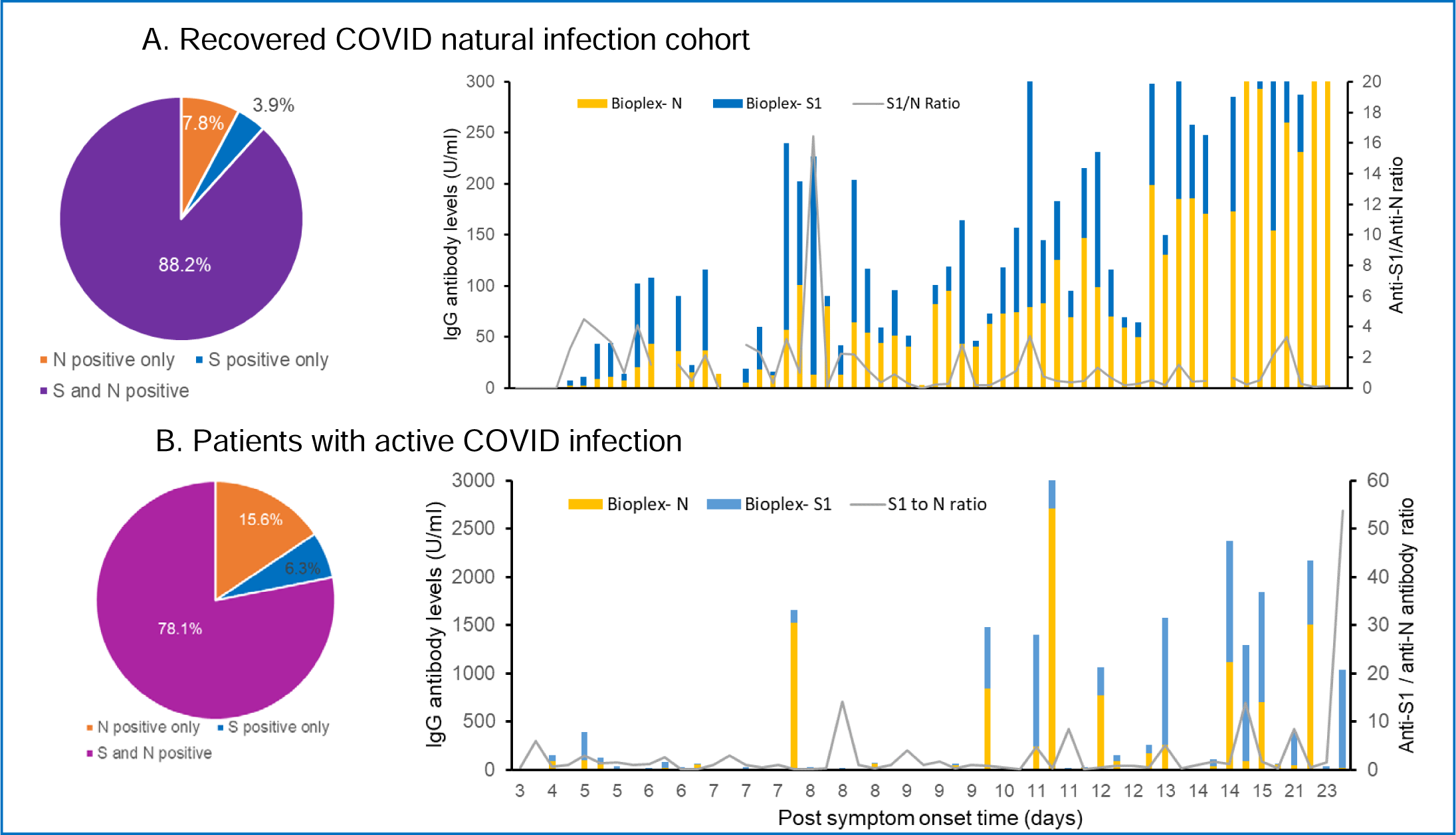
Imbalanced anti-S1 and anti-N antibody levels among naturally infected individuals. Anti-N and anti-S1 antibody levels within the same individual obtained using the Bioplex multiplex assay were compared. Most individuals developed both anti-S1 and anti-N antibodies after natural infection. Among them, 43.2% (19 of 44) in the recovery cohort showed a higher level of anti-S1 than anti-N antibodies. Anti-S1/anti-N ratio ranging from 0.07-16.26 with median =0.62. In the active infection group, 52.0% (13 of 25) showed a higher level of anti-S1 than anti-N antibodies. Anti-S1/anti-N ratio ranging from 0.05-53.74 with median =0.80. Between the recovery and active infection group, no significant difference was observed in anti-N antibody levels, anti-S1 antibody levels, or anti-S1/anti-N antibody ratio (p>0.05).

We also evaluated the correlation between antibodies against different protein targets in the acute COVID patients (Figure 3). Within the Bioplex panel, the anti-RBD and anti-S1 IgG levels showed a good positive correlation. This is expected, since RBD is part of the S1 protein. Between assays, the Atellica and Bioplex anti-RBD IgG antibody levels for the same sample also showed a strong positive correlation. However, neither anti-S2 nor anti-N antibody expression was positively correlated with anti-S1 production. On the contrary, the anti-S2 levels in the vaccinated cohort showed a strong positive correlation with anti-S1 levels (R2 = 0.9518). This difference may be attributed to the diversity in exposure, viral load, and viral-host interaction in natural infection versus the high consistency of vaccine dosage and its delivery.

**Fig 3.**
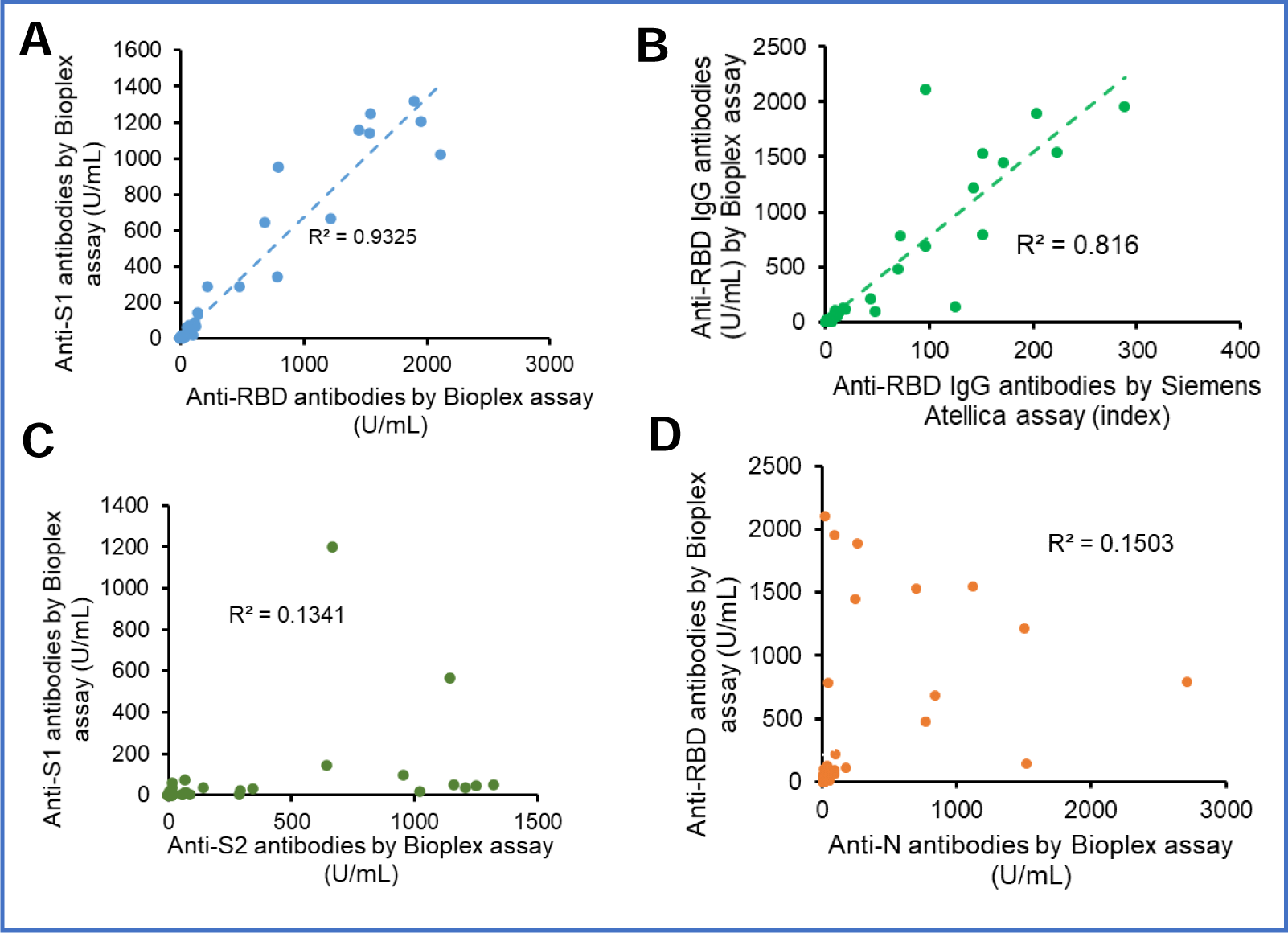
Correlation between SARA-CoV-2 antibodies against different antigens in individuals with acute natural infection. A: Anti-RBD antibody levels showed a strong positive correlation with anti-S1 antibody levels as expected. B: Anti-RBD antibody levels measured by different assays were also highly correlated. C: There was no positive correlation between the Anti-S2 and anti-S1 antibody levels. D: There was no positive correlation between the anti-N and anti-S1 antibody levels.

## Discussion

The study measured antibodies against various antigen targets and compared between three cohorts: 2-dose vaccinated HCW, unvaccinated HCW who recovered from natural infection, and patients during active COVID-19 disease. Our data demonstrated distinct differences in antibody profile following natural COVID infection and vaccination. Most infected individuals developed both anti-N and anti-RBD/S1 antibodies with greatly varied concentrations and about 54% have more anti-N than anti-S1 antibodies. Natural infection only induced a low level of anti-S2 antibodies in less than 50% of cases. All fully vaccinated individuals we tested showed strong immune reactivity toward RBD/S1, with an average of 50-fold higher antibody levels than naturally infected unvaccinated individuals. A response to S2 was also induced in all fully vaccinated subjects and the median value was approximately a 6-fold difference from that of a natural infection.

Compared to COVID-19 infection, a fundamental difference in the immune responses generated by the mRNA vaccines is antigen target specificity. Unlike a natural infection, where whole SARS-CoV-2 viruses elicit a large diversity of antibodies, the two FDA-approved mRNA vaccines only introduce the spike protein to the immune system. As evidenced by our study, the majority who recovered from COVID infection had both anti-N and anti-S antibodies with varied proportions, while vaccinated individuals without previous infection did not have detectable anti-N antibodies. The potential impact of the additional anti-N antibodies in naturally infected individuals remains undetermined. N protein is abundantly expressed during infection and highly immunogenic. It plays a key role in viral RNA replication and packaging into virions. Because the N protein is located within the viral envelope and not directly exposed to the host cells when the virus is intact, anti-N antibodies are unlikely to be neutralizing and not expected to play any role in disease prevention or reducing transmission. A study using isolated N-targeting mAbs from COVID-19 convalescents suggests some anti-N antibodies can inhibit complement hyperactivation and thus may lessen inflammation. Additionally, it is unclear whether the ratio between anti-N and anti-S is useful for predicting COVID-19 disease trajectories during the acute stage. Roltgen et al reported that higher ratios of IgG antibodies targeting S1 or RBD compared to N protein were seen in outpatients with mild illness versus severely ill patients (Roltgen et al. 2020). However, our result did not show any correlation between the anti-S1/anti-N ratio and the severity of the disease. This may due to the difference in antibody epitopes measured by assays. A study by Sen suggests that antibodies to a 21-residue epitope from nucleocapsid (Ep9) are associated with more severe disease outcomes (Sen et al. 2021).

A similar percentage of recovered and active infection subjects had only one type of detectable antibody (anti-S or anti-N), which suggests that those imbalanced antibody levels are unlikely due to differences in antibody decay. It rather indicates the variation in antibody production because of the difference in the interaction between the virus and host cells. Anti-N antibodies are often used as a serological marker for natural infection in areas where the vaccines only include or code spike proteins. Our data suggest a significant portion of the infected subjects who only produce anti-S antibodies might be missed when using anti-N for epidemiology study or investigation of COVID-19 associated Multisystem Inflammatory Syndrome in Children (MIS-C).

The S protein comprises subunits S1 and S2. S1 harbors the RBD and N-terminal domain (NTD). Both the Pfizer-BioNTech and Moderna COVID-19 vaccine encode a prefusion-stabilized, membrane-anchored SARS-CoV-2 full-length spike protein (Baden et al. 2021; Polack et al. 2020), and thus are capable of eliciting anti-RBD, S1, and S2 antibodies. Anti-RBD or anti-S1 antibodies are the major sources of neutralizing antibodies. Those antibodies might inhibit RBD binding to ACE2, thus preventing entry of SARS-CoV-2 into target cells. The 50-times higher S1/RBD antibody levels with vaccination relative to natural infection and the heterogeneity in antibody levels among the infected individuals strongly support that eligible individuals should consider vaccination regardless of previous COVID infection history. However, some unvaccinated but infected individuals may produce very high levels of anti-RBD/S1 antibodies that even exceed median vaccinated antibody levels. Additionally, the “hybrid immunity” (vaccinated after infection) exhibited 3- to 5-fold higher anti-RBD antibodies than the median level of naive vaccinated people. Those exceptionally high antibody levels may not necessarily be beneficial (Kojima and Klausner 2021; Nabin K. Shrestha 2021). With time, individual immunity to COVID becomes diversified due to frequency and levels of exposure, asymptomatic/symptomatic infection, and vaccination/booster doses. Individualized vaccination booster plans may need to be considered, at least for some special populations who have an unpredictable immune response in the future.

S2 harbors the fusion peptide domain, heptad repeat 1 (HR1), and HR2. It is necessary for the fusion of viral with host membranes. Anti-S2 antibodies are less studied. Theoretically, they may prevent membrane fusion, thus reducing viral entry. Additionally, S2 is structurally more conservative and less likely to harbor mutations than S1 (Shah et al. 2021). Antibodies against S2 may be a potential target for future therapeutic antibody and vaccine design for fighting existing and emerging variants. The lower levels of anti-S2 antibodies in both vaccinated and infected individuals compared to RBD and S1 antibodies may partially be due to the fact that the S2 subunit is not fully exposed until after receptor binding. The higher anti-S2 levels in the vaccinated versus infected cohort and strong positive correlation between anti-S2 and anti-S1/RBD antibody levels in vaccinated individuals indicate booster dose might be effective in bumping anti-S2 antibody production.

So far, it remains to be determined whether the serum anti-RBD or anti-S1 antibody levels can serve as an indicator for the degree of protection or the timing of booster administration. Neutralization antibody titer is considered the gold standard with a strong correlation to protective immunity, but the threshold level is not yet established. A breakthrough infection study in healthcare workers (Bergwerk et al. 2021) demonstrated that neutralizing antibody titers in cases of patients during the peri-infection period were lower than those in matched uninfected patients, and higher peri-infection neutralizing antibody titers were associated with lower infectivity (higher Ct values). Khoury et al used a mathematical model to analyze data from existing vaccine clinical trials and convalescent cohorts (Khoury et al. 2021). They estimated the neutralization level for 50% protection against detectable SARS-CoV-2 infection and severe disease to be 20.2% and 3% of the mean convalescent level, respectively. Recent studies show a varying degree of correlation between neutralizing antibody titers and anti-S1 or anti-RBD binding antibody values (Bal et al. 2021; Jaaskelainen et al. 2020). It should be noted that most of the assays in the existing studies are qualitative assays that may not have a linear response, especially at high antibody levels. The newer semi-quantitative assays, which typically have a much broader clinical reportable range necessary for measuring the high antibody concentration elicited by vaccination, have not been fully evaluated yet. However, current commercially available semi-quantitative assays use different units, and the numeric values can be dramatically different, even for the antibody against the same antigen target. Given the high correlation between assays for the same antigen targets, it is promising to standardize and correlate the anti-spike protein assays to neutralizing titer, and thus further evaluate their potential as a surrogate indicator for protective immunity and utility in vaccine booster planning.

A potential limitation of the study is that antibody activity does not encompass the entirety of the immune responses. T-cell immunity also plays an important role. Both COVID infection and vaccination can induce CD4 and CD8 T cell responses. Additionally, viral variants included in this study are predominantly the original SARS-CoV-2 strain and the strain with D614G mutation. Humoral response to infection with Delta or Omicron variant may be different.

In conclusion, individuals fully vaccinated with mRNA vaccine mounted strong humoral immunity with much higher anti-RBD1, anti-S1, and anti-S2 antibody levels compared to the naturally infected individuals. The results strongly support that all eligible individuals should be vaccinated, regardless of history of COVID-19 infection. Additionally, semi-quantitative anti-RBD/S assays that can quantify antibody levels with a broad reportable range are potentially useful in evaluating post-vaccination and/or post-infection immunity if a linear correlation with neutralizing antibody titers is achievable. Standardization and harmonization in quantitation and reporting are necessary for these assays to be useful in clinical practice.

## Data Availability

All data produced in the present study are available upon reasonable request to the authors.

## Acknowledgments

We thank Bio-Rad Laboratories and Siemens for providing reagents. We thank Behiye Goksel, YiYuan Zhou, and Caryn Good for help with sample collection and Rebecca Pontoni for assisting with antibody testing.

## Funding

This research received no specific grant.

**Supplementary Table S1.**
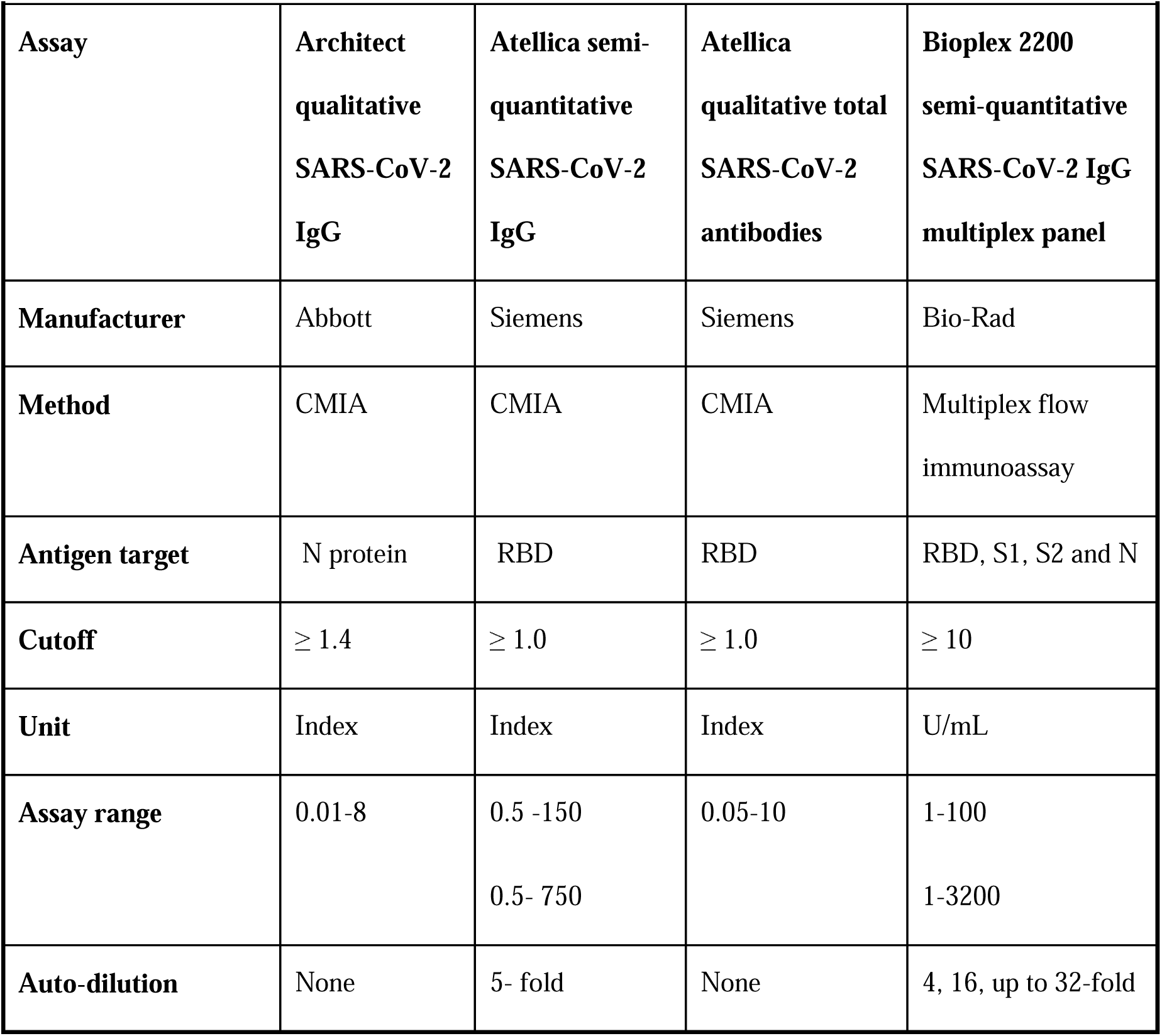
Description of SARS-CoV-2 antibody assays used in the study.

## Notes

### Competing Interest Statement

The authors have declared no competing interest.

### Funding Statement

This study did not receive any funding.

### Author Declarations

The study is part of a clinical laboratory method validation which has been reviewed by the Institutional Review Board of University Hospitals Cleveland Medical Center and determined as Non-Human Research and waived ethical approval.

## References

Baden LR, El Sahly HM, Essink B et al. (2021) Efficacy and Safety of the mRNA-1273 SARS-CoV-2 Vaccine The New England journal of medicine 384:403–416 doi:10.1056/NEJMoa2035389

Bal A, Pozzetto B, Trabaud MA et al. (2021) Evaluation of High-Throughput SARS-CoV-2 Serological Assays in a Longitudinal Cohort of Patients with Mild COVID-19: Clinical Sensitivity, Specificity, and Association with Virus Neutralization Test Clinical chemistry 67:742–752 doi:10.1093/clinchem/hvaa336

Bergwerk M, Gonen T, Lustig Y et al. (2021) Covid-19 Breakthrough Infections in Vaccinated Health Care Workers The New England journal of medicine 385:1474–1484 doi:10.1056/NEJMoa2109072

Bozio CH, Grannis SJ, Naleway AL et al. (2021) Laboratory-Confirmed COVID-19 Among Adults Hospitalized with COVID-19-Like Illness with Infection-Induced or mRNA Vaccine-Induced SARS-CoV-2 Immunity - Nine States, January-September 2021 MMWR Morbidity and mortality weekly report 70:1539–1544 doi:10.15585/mmwr.mm7044e1

Fowlkes A, Gaglani M, Groover K et al. (2021) Effectiveness of COVID-19 Vaccines in Preventing SARS-CoV-2 Infection Among Frontline Workers Before and During B.1.617.2 (Delta) Variant Predominance - Eight U.S. Locations, December 2020-August 2021 MMWR Morbidity and mortality weekly report 70:1167–1169 doi:10.15585/mmwr.mm7034e4

Hall VJ, Foulkes S, Charlett A et al. (2021) SARS-CoV-2 infection rates of antibody-positive compared with antibody-negative health-care workers in England: a large, multicentre, prospective cohort study (SIREN) Lancet 397:1459–1469 doi:10.1016/S0140-6736(21)00675-9

Jaaskelainen AJ, Kuivanen S, Kekalainen E et al. (2020) Performance of six SARS-CoV-2 immunoassays in comparison with microneutralisation Journal of clinical virology : the official publication of the Pan American Society for Clinical Virology 129:104512 doi:10.1016/j.jcv.2020.104512

Khoury DS, Cromer D, Reynaldi A et al. (2021) Neutralizing antibody levels are highly predictive of immune protection from symptomatic SARS-CoV-2 infection Nature medicine 27:1205–1211 doi:10.1038/s41591-021-01377-8

Kojima N, Klausner JD (2021) Protective immunity after recovery from SARS-CoV-2 infection The Lancet Infectious diseases doi:10.1016/S1473-3099(21)00676-9

Lopez Bernal J, Andrews N, Gower C et al. (2021) Effectiveness of the Pfizer-BioNTech and Oxford-AstraZeneca vaccines on covid-19 related symptoms, hospital admissions, and mortality in older adults in England: test negative case-control study Bmj 373: n1088 doi:10.1136/bmj.n1088

Nabin K. Shrestha PCB, Amy S. Nowacki, Paul Terpeluk, Steven M. Gordon (2021) Necessity of COVID-19 vaccination in previously infected individuals MedRxiv preprint doi: https://doi.org/10.1101/2021.06.01.21258176

Polack FP, Thomas SJ, Kitchin N et al. (2020) Safety and Efficacy of the BNT162b2 mRNA Covid-19 Vaccine The New England journal of medicine 383:2603–2615 doi:10.1056/NEJMoa2034577

Roltgen K, Powell AE, Wirz OF et al. (2020) Defining the features and duration of antibody responses to SARS-CoV-2 infection associated with disease severity and outcome Science immunology 5 doi:10.1126/sciimmunol.abe0240

Sara Y Tartof JMS, Heidi Fischer, Vennis Hong, Bradley K Ackerson, Omesh N Ranasinghe, Timothy B Frankland, Oluwaseye A Ogun, Joann M Zamparo, Sharon Gray, Srinivas R Valluri, Kaije Pan, Frederick J Angulo, Luis Jodar, John M McLaughlin (2021) Effectiveness of mRNA BNT162b2 COVID-19 vaccine up to 6 months in a large integrated health system in the USA: a retrospective cohort study Lancet 398:10 doi:10.1016/S0140-6736(21)02183-8

Sen SR, Sanders EC, Gabriel KN et al. (2021) Predicting COVID-19 Severity with a Specific Nucleocapsid Antibody plus Disease Risk Factor Score mSphere 6 doi:10.1128/mSphere.00203-21

Shah P, Canziani GA, Carter EP et al. (2021) The Case for S2: The Potential Benefits of the S2 Subunit of the SARS-CoV-2 Spike Protein as an Immunogen in Fighting the COVID-19 Pandemic Frontiers in immunology 12:637651 doi:10.3389/fimmu.2021.637651

Sheehan MM, Reddy AJ, Rothberg MB (2021) Reinfection Rates Among Patients Who Previously Tested Positive for Coronavirus Disease 2019: A Retrospective Cohort Study Clinical infectious diseases : an official publication of the Infectious Diseases Society of America 73:1882–1886 doi:10.1093/cid/ciab234

Shenai MB, Rahme R, Noorchashm H (2021) Equivalency of Protection From Natural Immunity in COVID-19 Recovered Versus Fully Vaccinated Persons: A Systematic Review and Pooled Analysis Cureus 13:e19102 doi:10.7759/cureus.19102

Vitale J, Mumoli N, Clerici P et al. (2021) Assessment of SARS-CoV-2 Reinfection 1 Year After Primary Infection in a Population in Lombardy, Italy JAMA internal medicine 181:1407–1408 doi:10.1001/jamainternmed.2021.2959

